# RISK OF ON JOB NON COMPLIANCE TOWARDS VARIOUS COVID-19 STANDARD & TRANSMISSION BASED INFECTION PREVENTION & CONTROL MEASURES/PRECAUTIONS AMONG THE HEALTHCARE WORKS WORKING IN OPD SETTINGs OF PUBLIC SECTOR TERTIORY CARE HOSPITALS OF QUETTA BALOCHISTAN

**DOI:** 10.1101/2021.07.08.21260212

**Authors:** Muhammad Arif, Muhammad Abdullah, Ambreen Chaudhary, Zakir Hussain, Ehsan Larik

## Abstract

**Background:** COVID-19 Pandemic is still circulating within the human population and proving to be a deadlier disease with mortality rate ranging from 0.5 to 7%^8^. Since COVID-19 is a highly transmissible disease; there is always a probability for its out ward spread towards general public and community from the hospitals and healthcare facilities where they come to seek treatment.

**Methodology:** A prospective cohort study design was used, considering the limited available resources and time __ A total of **200** healthcare workers *(Including Doctors, Nurses, Para-Medical staff, Janitorial staff, Reception staff & Pharmacists)* working in the OPDs of the two major Public sector hospitals of Quetta were made part of this study. The study participants were selected using **simple random sampling technique**. The study participants from **“Hospital-A”** were first of all educated and trained on various COVID-19 IPC measures later on various COVID-19-IEC materials; written in simple Urdu language, were displayed clearly everywhere in the OPD. Similarly hand washing station along with Hand sanitizers/Soaps and surgical face masks were also made available free of cost for all the study participants of Hospital-A. More over the importance and effectiveness of COVID-19 IPC measures were continuously announced in the OPD gallery of Hospital-A, these announcements used Simple wording in local languages (i.e. Urdu, Pashto, Balochi & Brahvi). On the other hand in the OPD of **“Hospital-B”** no such interventions were made. The study participants of both the hospitals were followed for one month and observations like-which group showed more on-job non compliance towards various COVID-19 IPC measures were recorded. The data was recorded on daily bases ***(From 1***^***st***^ ***May-to-31***^***st***^ ***May 2021)*** after observing the study participants and checklist was used for recording various findings. Lastly all the data was analyzed using Microsoft Excel 2007 version.

**Results:** The major findings of this study are almost in line with the set objectives, the study results are clearly showing the Risk ratio (R.R) as 0.27, indicating that the intervention group participants were only 27% as likely to develop On-job non-compliance for various COVID-19 IPC measures compare to the non-intervention group.

**Conclusion:** The best suggestion and intervention for the developing countries that could at least address the spread of COVID-19 in a cost effective manner at health facility levels remains to be adoption of various Standard and Transmission based non-pharmacological measures of Infection prevention and Control (IPC)^5^.

## Introduction

COVID-19 Pandemic is still circulating within the human population and proving to be a deadlier disease with mortality rate ranging from 0.5 to 7%^8^. According to the latest global data on WHO website as of **5:54pm CEST, 5 July 2021**, there have been **183**,**560**,**151 confirmed cases** of COVID-19, including **3**,**978**,**581 deaths** so far, as reported to WHO^1^. Due to lack of resources, surveillance and response structure in various developing countries; including Pakistan, less numbers of cases^3^ are reported daily.

As of **5 July 2021**, a total of **2**,**988**,**941**,**529 vaccine doses** have been administered^1^.Considering various factors like; low herd immunity, weak immunization pace, huge population numbers, poverty, illiteracy, weak health infrastructures and no vaccine production and research capabilities, most of the developing counties including Pakistan will continue to remain at risk for COVID-19 outbreaks^3^ in coming future.

Therefore the most safe and cost effective intervention for the developing countries therefore remains adoption of Infection prevention and Control (IPC) measures^5^.

According to WHO various IPC measures are poorly implemented/adopted in the various healthcare facilities of developing countries as a result, the prevalence of Hospital acquired infections (HCAIs) in low and middle-income countries varies between 5.7% and 19.1%. Similarly the proportion of patients with infections acquired in intensive care units in low and middle-income countries ranges from 4.4% to 88.9%, while the incidence of surgical site infections is up to nine times higher than in developed countries^24^.

Considering these points WHO came up with a universal Practical IPC guide lines for health care facilities in 2014^24^. This has been later on adopted and tailored by the health ministries of several low and middle-income countries including Pakistan.

### Literature Review

Most of infection prevention and control measures designed for healthcare facilities are mostly not disease centric in their approach, they are developed mostly by keeping two disease combating components into consideration **a)** Where is the reservoir of the disease causing bug **b)** what is the route of infection of that bug in the human body^5^.

Infection prevention and control measures for COVID-19 are classified into two categories ***<u>1)</u>*** *<u>Standard Precautions 2) Transmission base Precautions</u>*^24^. The standard precautions are mainly researched, designed and developed for the health care workers in mind because they are the most vulnerable group of people who has regular and close contacts with random patients in a hospitalized environment especially during OPD settings^5, 7, 8^.

The Standard precautions if adopted properly and regularly will block the entry of SARS-COV2 virus into the body of health care workers^17^.

The standard Precautions chock the entry points of SARS-COV2 virus which includes nose, mouth and eyes^10^.All the personal protective equipments (PPEs) are basically designed for this purpose hence health care workers are encouraged to wear face masks__ which will block the entry of SARS-COV2 virus droplets & particles through mouth and nose. Goggles__ for the eyes basically block the entry of suspended SARS-COV2 particles into the eyes. While Face shield__ gives additional cover over all the entry points of SARS-COV2 virus i.e. it covers all the entry points for the SARS-COV2 virus which includes eyes, nose & mouth^3, 11,12,13,24^. Other PPEs which includes Gown, Gloves, Head cover and Shoe cover are basically protecting the human skin from harboring SARS-COV2 virus on it; which can live for max of 7 days out of the human body while at minimum its life span is 4 hours^18, 19^, hence protecting against Indirect Infection through hands and surfaces if touched while dealing with a patient.

The Transmission based precautions for COVID-19 are helps to stop and minimize the COVID-19 transmission in a hospitalized environment; they include adopting Triage system, Regular hand washing, sanitization and disinfection protocols^7, 14, 22^.

So far several Information, education & communication (IEC) materials regarding and Behavioral change communication (BCC) materials have developed for COVID-19 but their real time effectiveness has not been studied yet.

### Operational Definitions

- <u>On Job:</u> “During duty hours in the hospital”.
- <u>Infection Prevention & control (IPC) measures:</u> “Refers to measures/activities intended to curtail infection and their spread in the healthcare facilities”^24^.
- <u>Information, education & Communication (IEC) material:</u> “Written/printed material in form of charts, broachers & pamphlets containing health education and health promotion knowledge used basically to improve the readers knowledge”^24^.
- <u>Behavioral change communication (BCC) materials:</u> **“**Refers to use of communication skills to promote positive health behaviors among the people communicated”^3^.
- <u>On Job Non compliance towards various COVID-19 IPC measures:</u> “Refers to not adopting or following the various COVID-19 IPC measures properly or as per protocol <u>or</u> Careless in adopting certain IPC measures during duty hours”^24^.

### Problem statement

COVID-19 is a highly transmissible disease; there is always a probability for its out ward spread towards general public and community from the hospitals and healthcare facilities where they come to seek treatment.

### Rationale

In developing countries like Pakistan where the prevalence of Hospital acquired infections (HCAIs) varies between 5.7% and 19.1% there is always a probability of SARS-COV2 virus infection. Normally the healthcare facilities in developing countries mostly belong to public sector where large number of patients visit daily to their OPD departments and seek treatments, therefore there is always a great possibility that these hospitals and healthcare facilities could harbor SARS-COV2 virus where it could mutate into a new potent stain (bug) that might cause an outward spill of towards the general public.

So far no published literature has assessed on job compliance towards standard and transmission based COVID-19 precautions among the healthcare workers working in a tertiary care hospital of a developing country especially after IEC and BCC interventions.

### Objectives

- To assess the incidence rate of On-job non compliance of various COVID-19 IPC measures among the two groups *(IPC trained group & untrained group)* of health care workers working in an OPD setup of a tertiary care hospital.
- To assess the impact of various COVID-19 trainings, IEC and BCC interventions on the attitude of the health care workers.

### Methodology

**A** prospective cohort study design was used, considering the limited available resources and time __ A total of **200** healthcare workers *(Including Doctors, Nurses, Para-Medical staff, Janitorial staff, Reception staff & Pharmacists)* working in the OPDs of the two major Public sector hospitals of Quetta were made part of this study. The study participants were selected using **simple random sampling technique**. The study participants from **“Hospital-A”** were first of all educated and trained on various COVID-19 IPC measures later on various COVID-19-IEC materials; written in simple Urdu language, were displayed clearly everywhere in the OPD. Similarly hand washing station along with Hand sanitizers/Soaps and surgical face masks were also made available free of cost for all the study participants of **Hospital-A**. More over the importance and effectiveness of COVID-19 IPC measures were continuously announced in the OPD gallery of Hospital-A, these announcements used Simple wording in local languages (i.e. Urdu, Pashto, Balochi & Brahvi). On the other hand in the OPD of **“Hospital-B”** no such interventions were made. The study participants of both the hospitals were followed for one month and observations like-which group showed more on-job non compliance towards various COVID-19 IPC measures were recorded. The data was recorded on daily bases ***(From 1***^**st**^ ***May-to-31***^**st**^ ***May 2021)*** after observing the study participants and checklist was used for recording various findings. Lastly all the data was analyzed using Microsoft Excel 2007 version.

## Results

### Descriptive statistics

**1. Socio-demographic Characteristics of Respondents:**

The major Socio-demographic characteristics of the Participants are summarized in the following table.

**Table #01:** Socio-demographic Characteristics of Respondents.

| S.NO | Socio-demographic Characteristics of the study participants | Sub-Set | Percentages (%) | Frequency (n) |
| --- | --- | --- | --- | --- |
| 1 | Education | Primary | 9% | 18 |
|  |  | Metric | 20% | 40 |
|  |  | Intermediate | 17% | 34 |
|  |  | MBBS/Graduation | 3% | 06 |
|  |  | Masters | 2% | 04 |
|  |  | Post graduation/Phd | 49% | 98 |
| 2 | Co-Morbidities | No Comorbidity | 90% | 180 |
|  |  | DM | 2% | 04 |
|  |  | Asthma | 4% | 08 |
|  |  | HTN | 2% | 04 |
|  |  | Hepatitis | 2% | 04 |
|  |  | Ovarian CA | 0% | 00 |
| 4 | COVID-19 Vaccination status | Vaccination No | 63% | 126 |
|  |  | Vaccination Yes | 37% | 74 |
|  | Hepatitis-B Vaccination Status | Vaccination Yes | 93% | 186 |
|  |  | Vaccination No | 7% | 14 |
| 5 | Seasonal flu Vaccination status | Seasonal Flu vaccination No | 7% | 14 |
|  |  | Seasonal Flu vaccination Yes | 93% | 186 |
| 6 | Blood Groups of the study participants | A+ | 7% | 14 |
|  |  | A- | 4% | 08 |
|  |  | B+ | 27% | 54 |
|  |  | B- | 3% | 06 |
|  |  | AB+ | 2% | 04 |
|  |  | AB- | 1% | 02 |
|  |  | O+ | 1% | 02 |
|  |  | O- | 1% | 02 |
|  |  | unkown | 54% | 108 |

**2. ALTERNATE DAY CHANGE IN THE ON JOB NON-COMPLIANCE ATTITUDE TOWARDS COVID-19 (IPC) MEASURES AMONG THE HEALTHCARE WORKERS OF THE <u>HOSPITAL-A</u>** *<u>(THE INTERVENTION GROUP):</u>*

On the very next day after the interventions in Hospital-A, it was observed that out of 100 study participants only 40% (n=40) were found to show full compliance towards all the COVID-19 IPC measures during their duty in the OPD. Post intervention observed on-job Compliance Level towards various COVID-19 IPC measures is shown below:

**Table # 02:** Post intervention observed on-job Compliance Level towards various COVID-19 IPC measures (Intervention group).

| **ALTERNATE DAY CHANGE IN THE <u>ON JOB NON-COMPLIANT ATTITUDE</u> TOWARDS COVID-19 (IPC) MEASURES AMONG THE HEALTHCARE WORKERS OF THE HOSPITAL-A (THE INTERVENTION GROUP)** |  |  |  |
| --- | --- | --- | --- |
| **S.NO** | **Date** | **Frequency of participants observed showing non compliance towards COVID-19 IPC Measures** | **Alternate day percent change in the non compliance level** |
| 1 | 1-May-21 | 60 |  |
| 2 | 2-May-21 | SUNDAY |  |
| 3 | 3-May-21 | 59 | #VALUE! |
| 4 | 4-May-21 | 55 | -7% ▼ |
| 5 | 5-May-21 | 55 |  |
| 6 | 6-May-21 | 55 |  |
| 7 | 7-May-21 | 53 | -4% ▼ |
| 8 | 8-May-21 | 52 | -2% ▼ |
| 9 | 9-May-21 | SUNDAY |  |
| 10 | 10-May-21 | 50 | #VALUE! |
| 11 | 11-May-21 | 50 |  |
| 12 | 12-May-21 | 50 |  |
| 13 | 13-May-21 | 49 | -2% ▼ |
| 14 | 14-May-21 | 47 | -4% ▼ |
| 15 | 15-May-21 | 45 | -4% ▼ |
| 16 | 16-May-21 | SUNDAY |  |
| 17 | 17-May-21 | 43 | #VALUE! |
| 18 | 18-May-21 | 43 |  |
| 19 | 19-May-21 | 43 |  |
| 20 | 20-May-21 | 40 | -7% ▼ |
| 21 | 21-May-21 | 40 |  |
| 22 | 22-May-21 | 40 |  |
| 23 | 23-May-21 | SUNDAY |  |
| 24 | 24-May-21 | 35 | #VALUE! |
| 25 | 25-May-21 | 33 | -6% ▼ |
| 26 | 26-May-21 | 32 | -3% ▼ |
| 27 | 27-May-21 | 30 | -6% ▼ |
| 28 | 28-May-21 | 27 | -10% ▼ |
| 29 | 29-May-21 | 27 |  |
| 30 | 30-May-21 | SUNDAY |  |
| 31 | 31-May-21 | 25 | #VALUE! |

**3. ALTERNATE DAY CHANGE IN THE ON JOB NON-COMPLIANCE ATTITUDE TOWARDS COVID-19 (IPC) MEASURES AMONG THE HEALTHCARE WORKERS OF THE <u>HOSPITAL-B</u>** *<u>(THE non-INTERVENTION GROUP):</u>*

From the very first day of observing non-compliant attitude among the 100 study participants of the Hospital-B (the non intervention group) were all poorly adopting the various COVID-19 IPC measures as shown below:

**Table # 02:** Observed on-job Compliance Level towards various COVID-19 IPC measures (Non-Intervention group).

| **ALTERNATE DAY CHANGE IN THE ON JOB NON-COMPLIANT ATTITUDE TOWARDS COVID-19 (IPC) MEASURES AMONG THE HEALTHCARE WORKERS OF THE HOSPITAL-B (THE NON INTERVENTION GROUP)** |  |  |  |
| --- | --- | --- | --- |
| **S.NO** | **Date** | **Frequency of participants observed showing non compliance towards COVID-19 IPC Measures** | **Alternate day percent change in the non compliance level** |
| 1 | 1-May-21 | 89 |  |
| 2 | 2-May-21 | SUNDAY |  |
| 3 | 3-May-21 | 92 | #VALUE! |
| 4 | 4-May-21 | 90 | -2% ▼ |
| 5 | 5-May-21 | 88 | -2% ▼ |
| 6 | 6-May-21 | 86 | -2% ▼ |
| 7 | 7-May-21 | 85 | -1% ▼ |
| 8 | 8-May-21 | 95 | 12% ▲ |
| 9 | 9-May-21 | SUNDAY |  |
| 10 | 10-May-21 | 92 | #VALUE! |
| 11 | 11-May-21 | 90 | -2% ▼ |
| 12 | 12-May-21 | 89 | -1% ▼ |
| 13 | 13-May-21 | 87 | -2% ▼ |
| 14 | 14-May-21 | 88 | 1% ▲ |
| 15 | 15-May-21 | 91 | 3% ▲ |
| 16 | 16-May-21 | SUNDAY |  |
| 17 | 17-May-21 | 88 | #VALUE! |
| 18 | 18-May-21 | 85 | -3% ▼ |
| 19 | 19-May-21 | 83 | -2% ▼ |
| 20 | 20-May-21 | 87 | 5% ▲ |
| 21 | 21-May-21 | 88 | 1% ▲ |
| 22 | 22-May-21 | 92 | 5% ▲ |
| 23 | 23-May-21 | SUNDAY |  |
| 24 | 24-May-21 | 91 | #VALUE! |
| 25 | 25-May-21 | 86 | -5% ▼ |
| 26 | 26-May-21 | 88 | 2% ▲ |
| 27 | 27-May-21 | 89 | 1% ▲ |
| 28 | 28-May-21 | 98 | 10% ▲ |
| 29 | 29-May-21 | 88 | -10% ▼ |
| 30 | 30-May-21 | SUNDAY |  |
| 31 | 31-May-21 | 90 | #VALUE! |

### Inferential Statistics

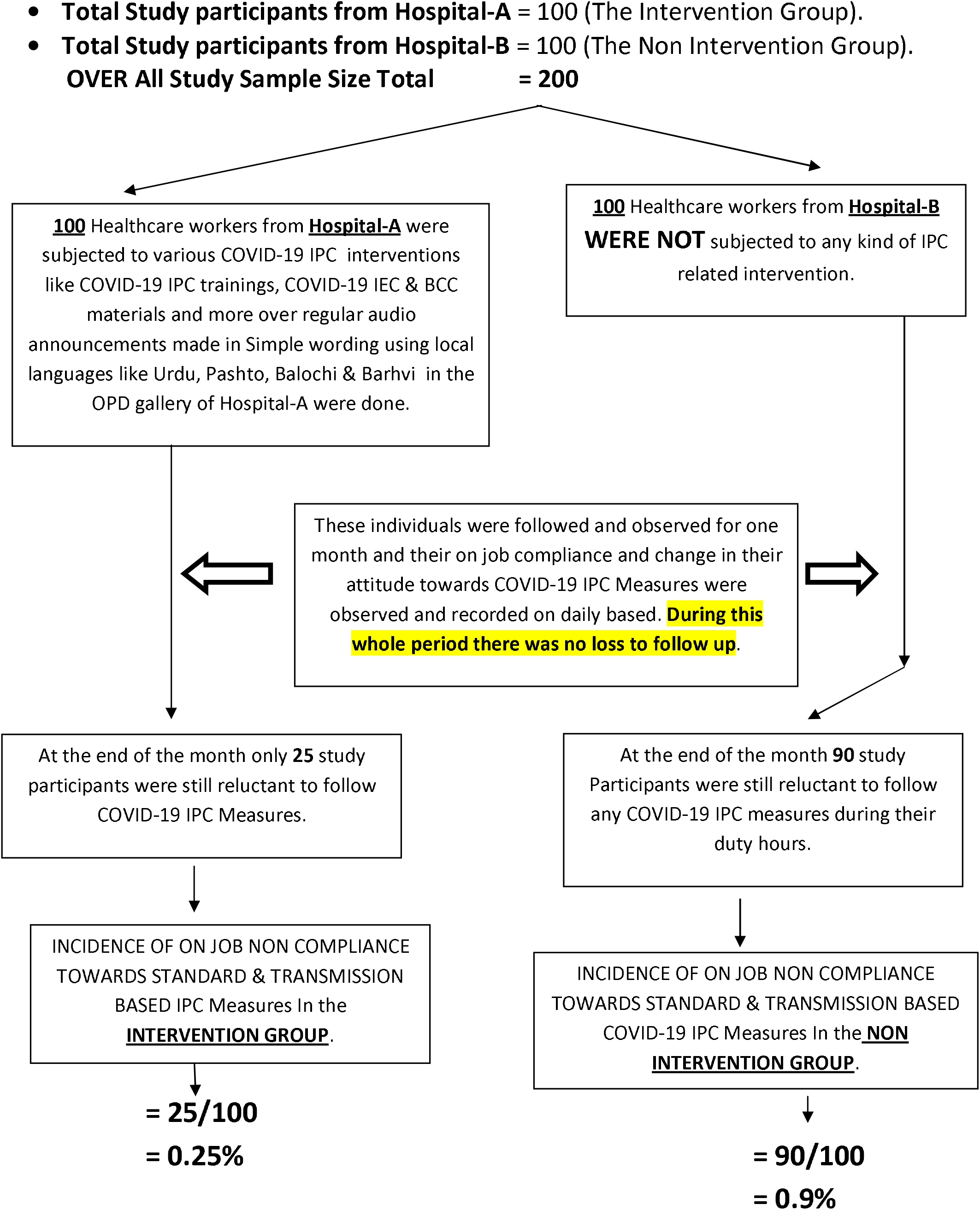

Using the incidence rates for non-compliance in both the groups the Risk Ratio Could be calculated as follow:

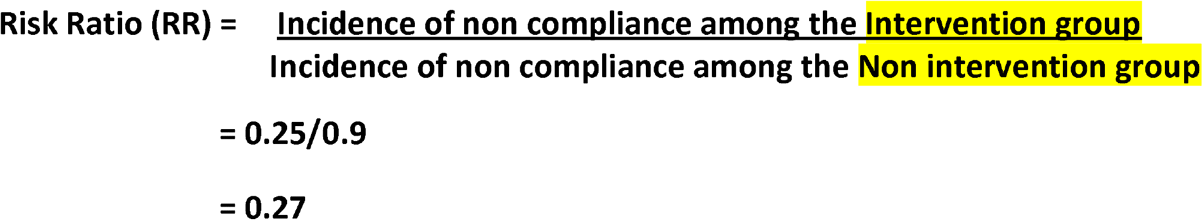

Hence those who were subjected to various COVID-19 IPC trainings and other interventions had <u>**0.27 times the risk**</u> for “*On job non-compliance towards various COVID-19 STANDARD & TRANSMISSION BASED Precautions*” compare to those who were not subjected to any intervention.

## Discussion & Conclusion

The major findings of this study are almost in line with the set objectives, the study results clearly showing that the **Risk ratio(R.R) of 0.27**, indicate that the intervention group participants were only 27% as likely to develop *On-job non-compliance* for various COVID-19 IPC measures compare to the non-intervention group. Considering the prevalence of Hospital acquired infections (HCAIs) in low and middle-income countries which vary between 5.7% and 19.1%^24^ there is always a high probability that these infections including COVID-19^3^ could spill out of the Hospitals environment into the community and also they can infect Healthcare workers working over there. Hence With low herd immunity, weak immunization pace, huge population numbers, poverty, illiteracy, weak health infrastructures, no vaccine production and research capabilities most of the developing counties including Pakistan will remain at risk for COVID-19 outbreaks in future.

The best suggestion and intervention for the developing countries that could at least address the spread of COVID-19 in a cost effective manner at health facility levels remains to be adoption of various Standard and Transmission based non-pharmacological measures of Infection prevention and Control (IPC)^5^

### Recommendations

It is highly recommended various COVID-19 specific infection prevention & control interventions like COVID-19 IPC trainings, COVID-19 IEC & BCC materials to be displayed clearly everywhere in the healthcare facilities especially in OPD department. Moreover audio announcements made in Simple wording using local languages like Urdu, Pashto, Balochi & Barhvi could really serve as constant reminder tools especially in a OPD department_ where every NEXT patient in queue could present with a different infectious bug.

## Data Availability

Data would be shared upon request.

